# Epigenetic and Genetic Profiling of Comorbidity Patterns among Substance Dependence Diagnoses

**DOI:** 10.1101/2024.10.08.24315111

**Authors:** Gita A. Pathak, Robert H. Pietrzak, AnnMarie Lacobelle, Cassie Overstreet, Frank R. Wendt, Joseph D. Deak, Eleni Friligkou, Yaira Nunez, Janitza L. Montalvo-Ortiz, Daniel F. Levey, Henry R. Kranzler, Joel Gelernter, Renato Polimanti

## Abstract

**Objective:** This study investigated the genetic and epigenetic mechanisms underlying the comorbidity patterns of five substance dependence diagnoses (SDs; alcohol, AD; cannabis, CaD; cocaine, CoD; opioid, OD; tobacco, TD).

**Methods:** A latent class analysis (LCA) was performed on 31,197 individuals (average age 42±11 years; 49% females) from six cohorts to identify comorbid DSM-IV SD patterns. In subsets of this sample, we tested SD-latent classes with respect to polygenic burden of psychiatric and behavioral traits and epigenome-wide changes in three population groups.

**Results:** An LCA identified four latent classes related to SD comorbidities: AD+TD, CoD+TD, AD+CoD+OD+TD (i.e., polysubstance use, PSU), and TD. In the epigenome-wide association analysis, *SPATA4* cg02833127 was associated with CoD+TD, AD+TD, and PSU latent classes. AD+TD latent class was also associated with CpG sites located on *ARID1B*, *NOTCH1*, *SERTAD4,* and *SIN3B*, while additional epigenome-wide significant associations with CoD+TD latent class were observed in *ANO6* and *MOV10* genes. PSU-latent class was also associated with a differentially methylated region in *LDB1*. We also observed shared polygenic score (PGS) associations for PSU, AD+TD, and CoD+TD latent classes (i.e., attention-deficit hyperactivity disorder, anxiety, educational attainment, and schizophrenia PGS). In contrast, TD-latent class was exclusively associated with posttraumatic stress disorder-PGS. Other specific associations were observed for PSU-latent class (subjective wellbeing-PGS and neuroticism-PGS) and AD+TD-latent class (bipolar disorder-PGS).

**Conclusions:** We identified shared and unique genetic and epigenetic mechanisms underlying SD comorbidity patterns. These findings highlight the importance of modeling the co-occurrence of SD diagnoses when investigating the molecular basis of addiction-related traits.

## INTRODUCTION

Substance dependence (SD) is a significant public health concern, affecting more than 40 million individuals in the United States ^1^. The most prevalent SDs are related to alcohol (AD), cannabis (CaD), cocaine (CoD), opioid (OD), and tobacco (TD). Patients with SDs often exhibit misuse of more than one substance, which complicates treatment and recovery efforts ^2^. The comorbidity of SDs exacerbates a range of negative health outcomes, making it critical to understand the patterns of SD comorbidities ^2, 3^. The heterogeneity of SD comorbidity patterns poses a major challenge in identifying the mechanistic underpinnings of these disorders. SD patterns can stem from a range of factors, including shared genetic predisposition, environmental influences, and sociocultural context. Genome-wide association studies (GWAS) and epigenome-wide association studies (EWAS) uncovered mechanisms contributing to the predisposition to SDs ^4–10^. To date, these previous efforts largely focused on single SDs, limiting our understanding of the real world in which patients are generally affected by multiple SDs.

This study identified comorbidity patterns among AD, CaD, CoD, OD, and TD using a latent class analysis (LCA, a method that aims to identify more homogeneous subgroups in heterogeneous data). Then, we investigated SD-latent classes with respect to methylation changes and polygenic burdens related to psychiatric and behavioral traits (**Figure 1**). The findings shared and unique epigenetic and genetic profiles that underlie SD comorbidity patterns.

**Figure 1.**
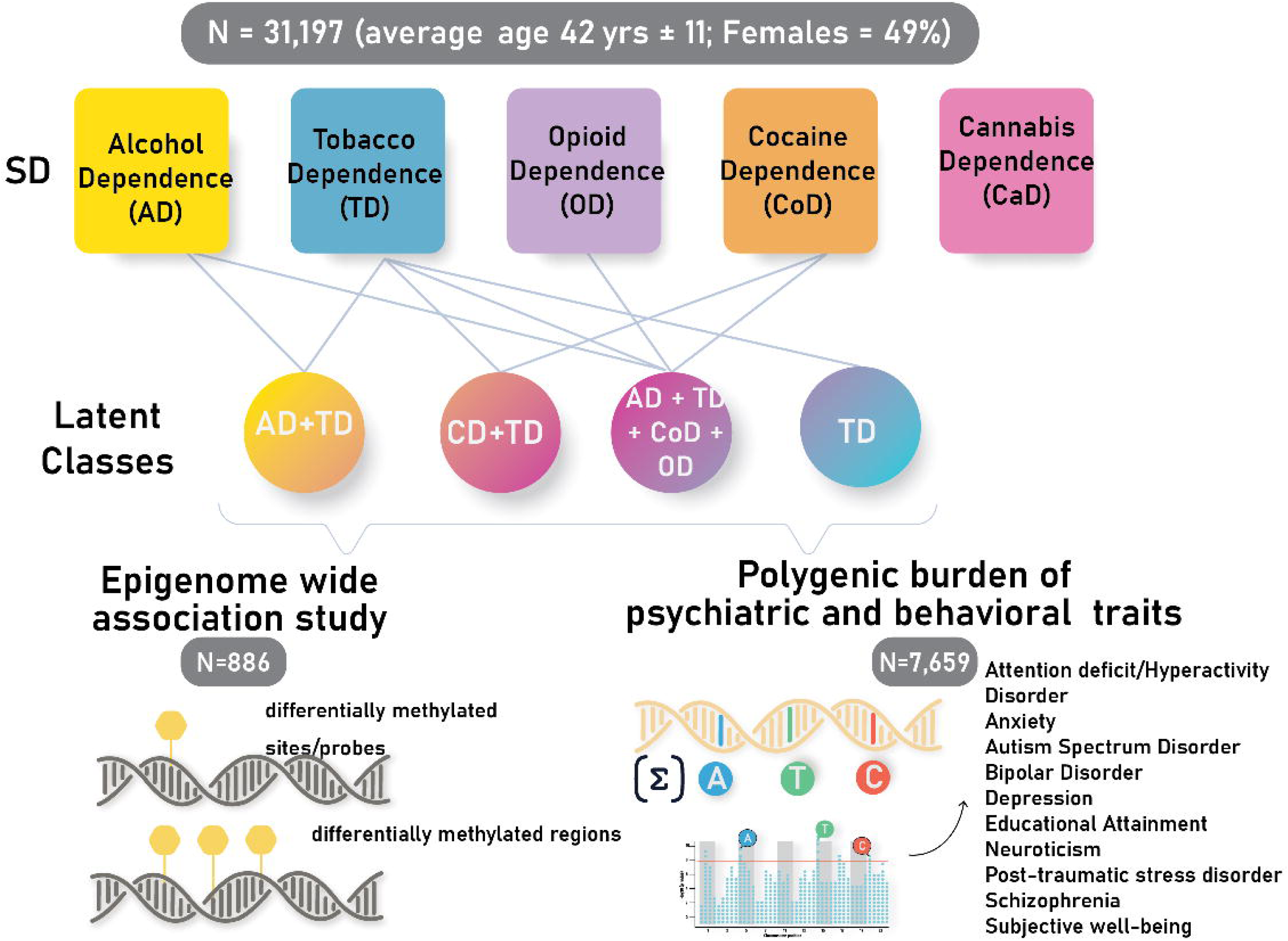
Study Design. An overview of the study design showing five substance dependence (SD) diagnoses assessed in six different cohorts and used to identify latent classes related to their comorbidities. Each of the SD latent classes was compared against the control group in the epigenome-wide association study and estimating genetic burden of ten psychiatric and behavioral traits.

## METHODS

### Cohorts

We investigated six cohorts with information regarding DSM-IV diagnoses of AD, CaD, CoUD, OD, and TD (**Table 1**). In addition to the Yale-Penn cohort^11–15^, we analyzed five datasets available from NCBI’s Database of Genotypes and Phenotypes (dbGaP): ‘Study of Addiction: Genetics and Environment’ [phs000092] ^16^, ‘Genome-Wide Association Study of Heroin Dependence’ [phs000277] ^17^, ‘Genetic Architecture of Smoking and Smoking Cessation’ [phs000404]^18^, ‘CIDR, NCI, NIDA Sequencing of Targeted Genomic Regions Associated with Smoking’ [phs000813] ^18^, and ‘Nicotine Addiction Genetics and Correlates’ [phs001299] ^19^. Overall, we analyzed SD data collected from 31,197 individuals (average age 42±11 years; 49% females).

**Table 1:** Characteristics of the cohorts investigated.

|  | **N** | **Age,**<br>**mean±SD** | **Female %** | **AD, N** | **CaD, N** | **CoD, N** | **OD, N** | **TD, N** |
| --- | --- | --- | --- | --- | --- | --- | --- | --- |
| **phs000092** | 4,121 | 39±9 | 54 | 1,946 | 753 | 1,130 | 304 | 1,856 |
| **phs000277** | 3,515 | 46±10 | 42 | 55 | 4 | 0 | 0 | 205 |
| **phs000404** | 1,527 | 37±6 | 59 | 204 | 222 | 149 | 21 | 1,239 |
| **phs000813** | 2,969 | 52±7 | 60 | 516 | 354 | 210 | 57 | 1,844 |
| **phs001299** | 3,508 | 50±14 | 51 | 493 | 134 | 6 | 32 | 1,367 |
| **Yale-Penn** | 15,557 | 40±12 | 46 | 7,481 | 3,897 | 8,662 | 4,379 | 8,219 |
| **Total** | 31,197 | 42±11 | 49 | 10,695 | 5,364 | 10,157 | 4,793 | 14,730 |

### Latent Class Analysis

LCA was performed in R version 4.1 using the poLCA R package ^20^for the primary investigation and Mplus software ^21^for validation. The number of latent classes was identified based on the lowest Akaike Information Criterion (AIC), Bayesian Information Criterion (BIC), Likelihood ratio/deviance statistic (G^2^), and Pearson Chi-square goodness of fit statistic (χ^2^). The probability threshold to assign participants with SDs in classes was identified using MANOVA by comparing the probability of SD cases across the latent classes identified in the best-fitting model.

### Genetic Data

Genotype data from the six studies was cleaned by removing individuals with mismatched biological sex, low genotyping rate, heterozygosity, and relatedness. Single nucleotide polymorphisms (SNPs) were removed based on minor allele frequency (MAF<1%), Hardy-Weinberg equilibrium (p<1×10^−6^), and sample missingness (<10%). Continental genetic ancestry was estimated against a combined reference panel including 1000 Genomes Project and Human Genome Diversity Project ^22^. Genetic relatedness and within-ancestry principal components were generated using KING ^23^. Genotype data from each study was imputed using TopMed Imputation server ^24^. Because large-scale psychiatric/behavioral GWAS are present only for populations of European descent, we limited the PGS analysis to this population group. After quality control (QC), genetic data from 7,659 individuals of European descent were available for PGS testing.

### Epigenetic Data

DNA methylation data were available for a subset of the Yale-Penn participants (n=886). Briefly, DNA was extracted from whole blood of Yale-Penn participants collected at the time of recruitment using Paxgene Blood DNA Kit (Qiagen, MD, USA). Bisulfite conversion of the extracted DNA was performed using the EZ-96 DNA methylation kit (Zymo Research, CA, USA) and subsequently processed on the Infinium ® MethylationEPIC BeadChip array (Illumina, San Diego, CA USA) which assesses the methylation status of over 850,000 CpG sites. Raw signal intensity data (IDAT) files from the array were uploaded and processed in R Studio. These files provided beta values, indicating DNA methylation levels at specific CpG sites, with values ranging from 0 (unmethylated) to 1 (fully methylated). Primary QC steps removed CpG sites with low detection p-values, missing beads, close proximity to SNPs, multi-hit sites, and non-autosomal sites. The beta values of the remaining CpG sites were normalized using the beta mixture quantile (BMIQ) method ^25^. Batch effects were identified using singular vector decomposition. Technical batch effects related to array and slide were corrected using ComBat function in sva R package ^26^. DNA Methylation data were used to estimate blood cell-type compositions (i.e., CD8+T, CD4+T, natural killer, B, monocytes, and neutrophils) ^27^, and smoking status ^28^.

### Epigenome-wide association analysis

After QC, we tested 657,226 CpG sites for differential methylation with respect to SD-latent classes in three population groups (European-descent, EUR, n=481; African-descent, AFR n=339; Admixed-Americans, AMR n=66). Association analysis for CpG sites was performed on M-values (transformed beta values) using empirical Bayes methods implemented in the limma R package ^29^. The analysis was adjusted for age, sex, genotype-derived principal components 1-10, methylation-based smoking score ^28^, and proportions of blood cell types (i.e., CD8+T, CD4+T, natural killer, B, monocytes, and neutrophils). Inflation was calculated using the QQperm R package ^30^. For the ancestry-stratified EWAS of each SD-latent class, the minimum sample size was 15. The meta-analysis across all ancestries was performed using GWAMA ^31^ to improve statistical power. For each EWAS cross-ancestry meta-analysis, genomic control correction was applied to epigenome-wide association statistics when lambda was >1.05. Differentially methylated regions were identified using the dmrff R package ^32^. Regions were defined as consisting of 2 to 30 CpG sites within 500 base pairs. False discovery rate (FDR q<0.05) was used to adjust for multiple testing. The CpG sites were further investigated using information available from the EWAS Catalogue^33^ (available at https://www.ewascatalog.org/). We also assessed brain-blood concordance of the CpG sites identified using BECon application ^34^.

### Polygenic Score Analysis

PGSs were derived from GWAS of attention deficit/hyperactivity disorder (ADHD; N=225,534) ^35^, anxiety (N=1,096,458) ^36^, autism spectrum disorder (N=46,350) ^37^, bipolar disorder (N=413,466) ^38^, depression (N=1,035,760) ^39^, educational attainment (N=765,283) ^40^, neuroticism (N=380,000) ^41^, posttraumatic stress disorder (PTSD; N=1,222,882) ^42^, schizophrenia (N=320,404) ^43^, and subjective well-being (N=298,420) ^44^. The polygenic scores were calculated using PRS-CS^45^, with the 1000 Genomes reference panel for linkage disequilibrium, scaled to have mean of 0 and a unit of 1 standard deviation. In each cohort, PGSs were tested with respect to SD-latent classes, including age, sex, and top-ten within-ancestry principal components as covariates. For each SD-latent class, cohort-specific PGS associations were meta-analyzed using meta R package^46^. Bonferroni-corrected threshold of p<1.25×10^−3^ was used to adjust for the number of PGS association tests performed (n=40).

## RESULTS

### Latent class analysis of SDs

With respect to comorbidity patterns among DSM-IV based AD, CaD, CoD, OD and TD, the model with five latent classes was identified as the best fitting because of the lowest value combination of AIC, BIC, G^2^, and χ^2^ metrics (**Supplementary Table 1**). Considering a posterior probability≥70% (**Figure 2**; **Supplementary Figure 1**), we stratified the sample of 31,197 subjects across the five SD-latent classes. Because of the high case-posterior probability for AD and TD (97% and 75%, respectively), 6,487 participants were included in the AD+TD-latent class. A total of 1,170 participants were assigned to the CoD+TD-latent class, because of the high case-posterior probability of these SDs (85% and 70%, respectively). We also observed 2,090 participants related to a polysubstance use disorder (PSU)-latent class, which had high case-posterior probabilities for AD (95%), CoD (99%), OD (100%), and TD (96%). An additional latent class including 1,162 individuals showed high posterior probability only for TD (100%). Finally, there were 11,759 participants assigned to a latent class with high control-posterior probability for all SDs (i.e., 89% AD, 80% CaD, 94% CoD, 72%TD, 99% OD). CaD did not show high case-posterior probability in any of the SD-latent classes (**Supplementary Table 2**).

**Figure 2.**
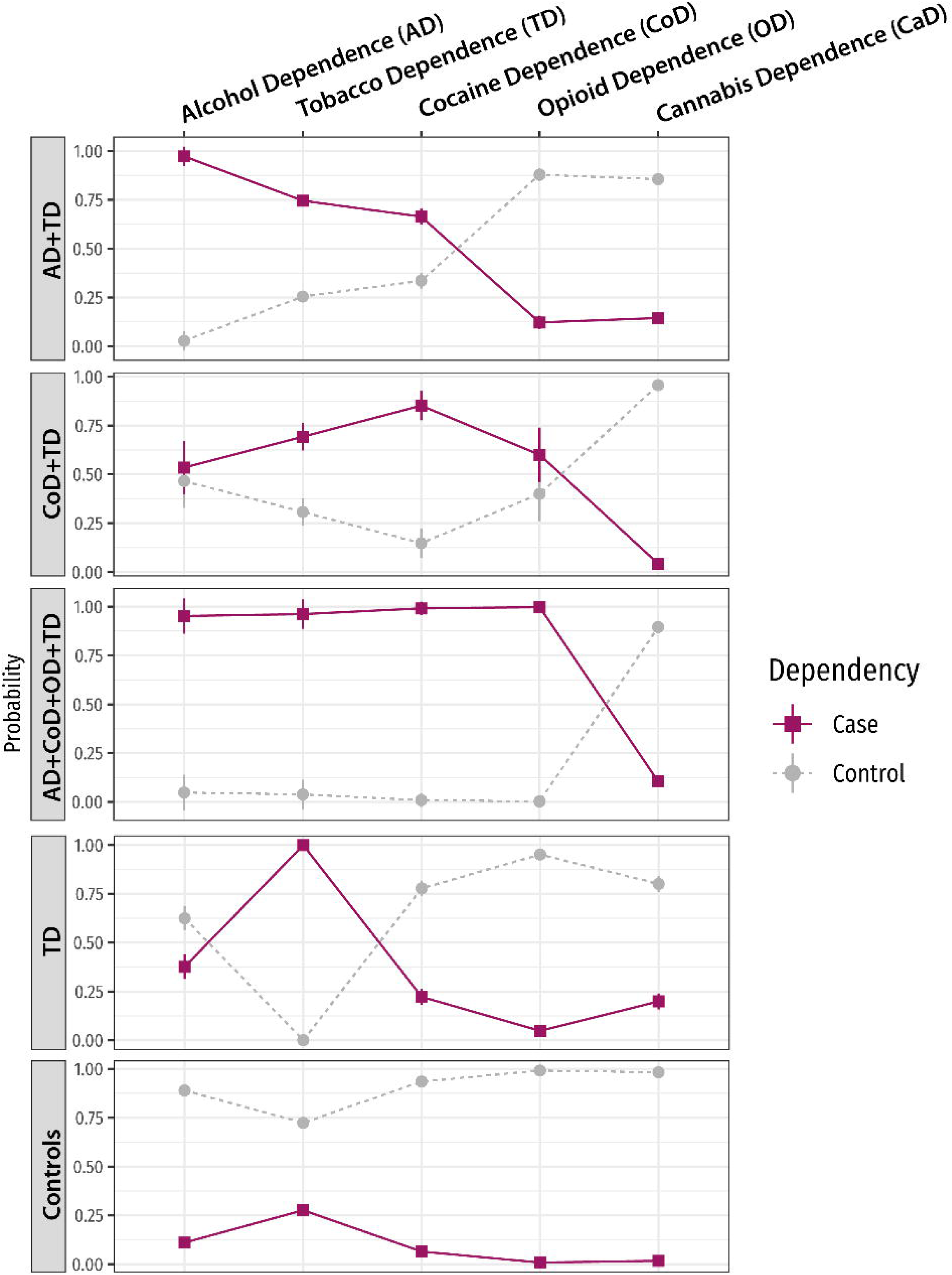
Distribution of case- and control-posterior probability across SDs in the five-latent class model. The x-axis (top) shows each of the substance dependence diagnoses, and the y-axis shows the probability value of cases and controls for each of the five SDs across each latent class shown on the left. Details of the statistical comparisons are shown in Supplementary Figure 1.

### Epigenome-wide association study of SD latent classes

Relative to the control-latent class group, we performed cross-ancestry EWAS meta-analyses of AD+TD, CoD+TD, PSU-latent classes in 886 individuals, identifying seven CpG associations (**Figure 3**; **Supplementary Figure 2; Supplementary Tables 3-6**). TD-latent class was excluded from this analysis because of the small sample size. After FDR multiple testing correction (FDR q<0.05), *SPATA4* cg02833127 (location: 1^st^ exon, 5’UTR) was associated with the three SD-latent classes investigated (i.e., AD+TD β=-0.49, P=9.9×10^−50^; CoD-TD β=-0.3, *P*= 2.93×10^−11^; PSU β=-0.24, *P*=3.19×10^−11^). Four additional CpG sites were associated with the AD+TD-latent class: *SIN3B* cg06815056 (location: TSS200; β=0.23, P=2.91×10^−7^), *ARID1B* cg19436567 (location: 1^st^ exon; β=0.22, P=1.47×10^−7^), *SERTAD4* cg20270863 (location: TSS200, 5’UTR; β=0.16, P=3.42×10^−7^) and *NOTCH1* cg13725899 (location: gene body; β=0.16, P=8.69×10^−8^). Two additional FDR-significant CpG associations were detected with respect to CoD+TD latent class: *MOV10* cg08355353 (Location gene body; β=0.43, *P*=1.08×10^−7^) and *ANO6* cg09909775 (Location: 1stExon, 5’UTR; β=0.50; P=2.03×10^−7^). Considering information available from the EWAS catalog (**Supplementary Table 7**), these CpG sites were previously identified in the context of aging (cg08355353, cg06815056, cg13725899, cg19436567, and cg02833127), rheumatoid arthritis (cg06815056), Alzheimer’s disease (cg13725899), renal carcinoma (cg20270863), and molecular regulation (cg06815056, cg13725899, and cg19436567).

**Fig 3.**
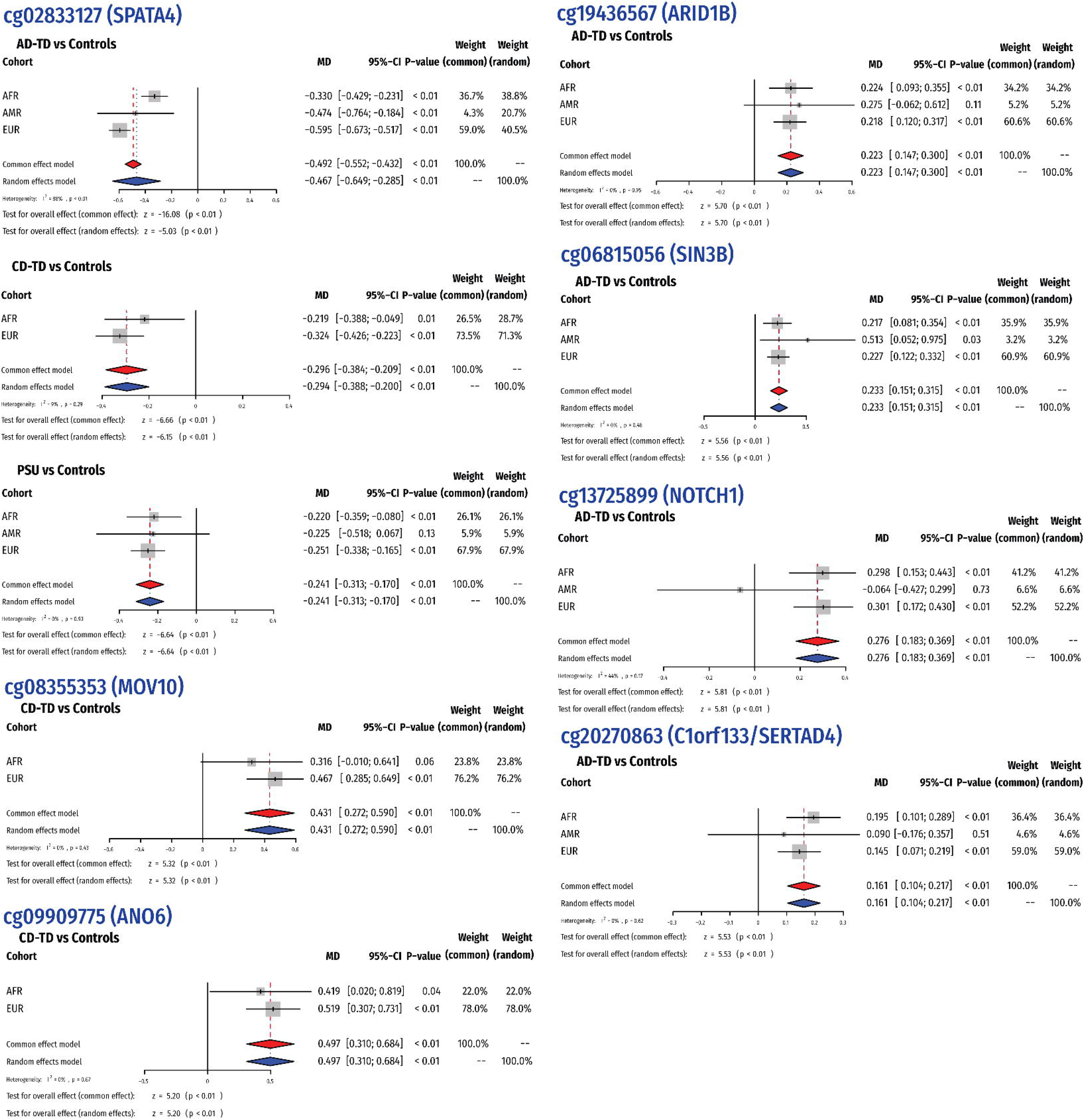
Epigenome-wide association study of SD latent classes. Forest Plots show meta-analyzed effect sizes of the significant CpG sites across ancestral groups; EUR – European descent; AFR – African descent; AMR – Admixed Americans.

We observed cross-ancestry heterogeneity in the association between *SPATA4* cg02833127 and AD+TD-latent class (I^2^=88%, heterogeneity p<2×10^−4^) where the effect was more negative in EUR (β = -0.60, 95%CI=-0.67 – -0.52) than in AFR (β = - 0.33, 95%CI=-0.43 – -0.23). Conversely, no cross-ancestry heterogeneity was observed in *SPATA4* cg02833127 association with CoD+TD and PSU-latent classes and also for the other FDR-significant CpG associations (heterogeneity-p>0.05; **Figure 3**).

In addition to CpG association, we also observed one FDR-significant differentially methylated region (chr10:102120371-102120478; *Z*=5.466; *P*= 4.60×10^−8^) related to PSU-latent class, spanning 2 CpG sites (cg15320455 – TSS200; cg17106419 – TSS200) within *LDB1* gene (**Supplementary Table 8**).

Comparing brain-blood methylation patterns of the identified CpG sites (**Supplementary Figure 2**), we observed correlation estimates in the top-90^th^ percentile for *MOV10* cg08355353 (rho=0.36 in the Brodmann area (BA) 7), *SERTAD4* cg20270863 (rho=0.38 in BA20), *SIN3B* cg06815056 (rho=-0.45 in BA20), *SPATA4* cg02833127 (rho=-0.44 in BA7), and *ARID1B* cg19436567 (rho=0.49 in BA10; rho=-0.35 in BA20)

### Polygenic burden associations with SD-latent classes

Assessing the pleiotropy across SD-latent classes, we observed different patterns of psychiatric-behavioral PGS associations (**Figure 4**; **Supplementary Table 9**). After Bonferroni multiple testing correction (p<1.25×10^−3^), PTSD was the only PGS associated across the four SD-latent classes (**Supplementary Table 9**). TD-latent class was only associated with PTSD PGS (Odds ratio, OR=1.32, p=7.24×10^−4^). In contrast, PSU, AD+TD, and CoD+TD latent classes showed shared PGS associations with ADHD (positive relationship), anxiety (positive relationship), educational attainment (inverse relationship), and schizophrenia (positive effect) with comparable effect sizes (**Supplementary Table 9**). In addition to shared pleiotropic mechanisms, we also observed specific PGS associations. PSU-latent class was uniquely associated with PGS of neuroticism (OR=1.21; P=2.62×10^−6^) and subjective wellbeing (OR=0.86; P=2.12×10^−4^) with opposite effect directions. AD+TD-latent class was uniquely associated with bipolar disorder-PGS (OR=1.14; P=1.39×10^−5^).

**Fig 4.**
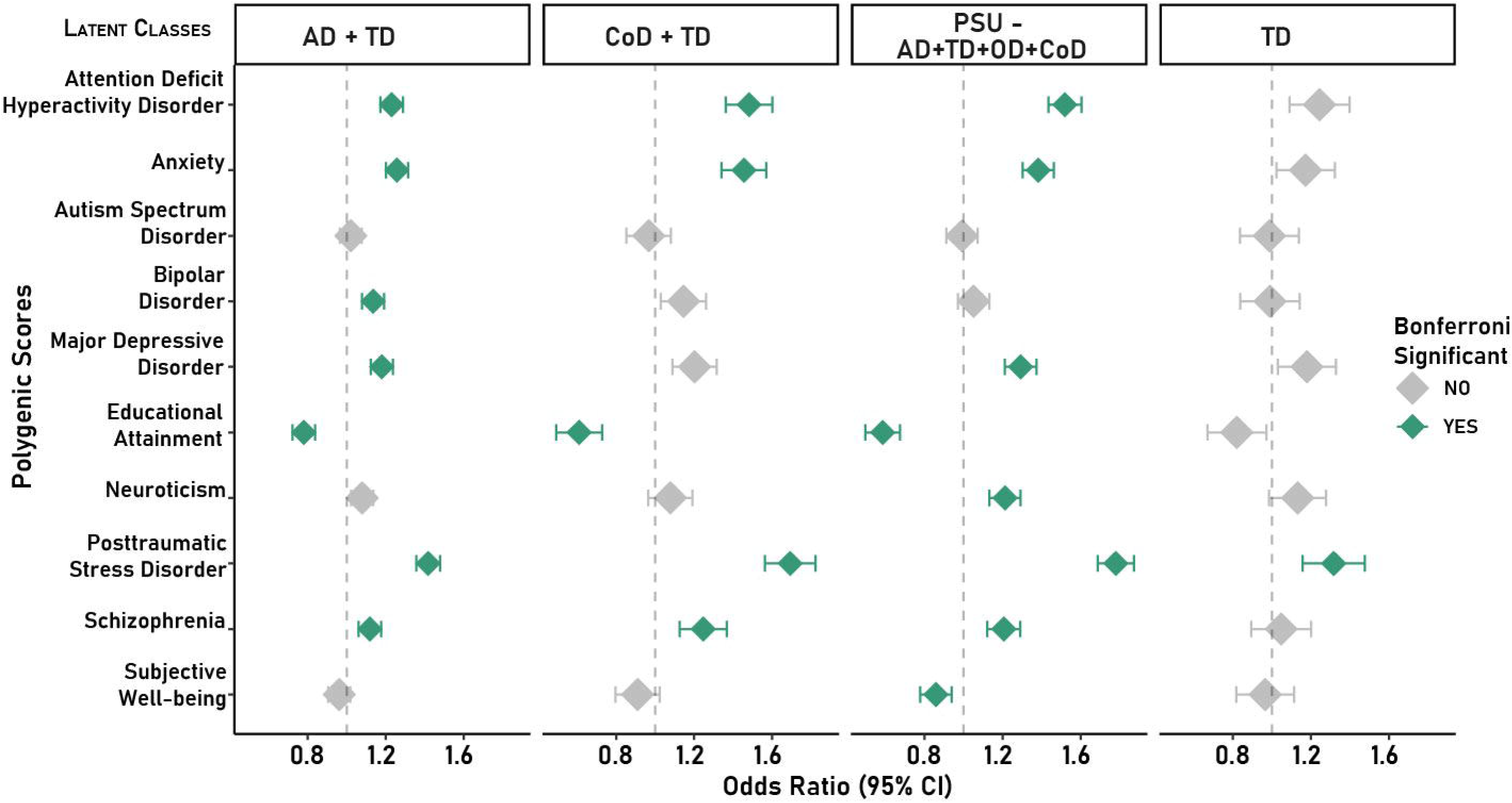
Association of polygenic scores of psychiatric and behavioral traits with SD latent classes. Forest plot showing meta-analyzed associations of ten traits with SUD latent classes. The x-axis shows odds ratio as points and 95% confidence intervals as lines. The Bonferroni-significant associations are marked with green, while others are indicated in grey. Details regarding PGS associations are shown in Supplementary Table 9.

Because PGS associations were estimated from the meta-analysis of cohorts with different characteristics (**Table 1**), we also estimated cross-cohort heterogeneity within the PGS effects. After Bonferroni correction, there was significant heterogeneity (heterogeneity Q-P<1.25×10^−3^) in the association between PTSD PGS and AD+TD-latent class (I2=0.89 ; tau^2^=0.05; Q=28.4, Q-df=3, Q-p=3×10^−6^) and between educational attainment PGS and TD-latent class (I2=0.84; tau^2^=0.12; Q=18.9, Q-df=2, Q-P=2.81×10^−4^). In the first, the heterogeneity was driven by two-studies, phs000404 and phs001299 (**Supplementary Tables 9and 10**). The meta-analyzed effect association between educational attainment PGS and TD-latent class was only nominally significant (OR=0.82, P=0.01) and its Bonferroni-significant heterogeneity was driven by two studies, Yale-Penn and phs000092 (**Supplementary Tables 9and 10**).

## DISCUSSION

SDs have a pervasive negative impact on individuals, families, and society at large. More severe consequences are experienced by individuals with co-occurring SDs, which represent the largest portion of affected individuals^47^. To date, most genomic studies modeled singular SD cases without considering co-occurring SDs ^9, 10^. Recently, large-scale GWAS datasets have been used to investigate the shared pathogenesis across SDs and related phenotypes^48, 49^. While the findings of these studies increased our understanding of the molecular mechanisms responsible for polysubstance comorbidities, more efforts are needed to dissect the dynamics underlying different patterns of SD co-occurrence. Applying LCA, previous observational studies of nationally representative samples, treatment-seeking populations, and cohorts enriched for SD cases identified latent classes reflecting different polysubstance patterns ^50–54^. Building on this evidence, we identified four SD-latent classes in a sample of >30,000 individuals and then investigated genetic and epigenetic mechanisms associated with the different SD comorbidity patterns.

Investigating epigenome-wide differences, our study revealed several differentially methylated sites with shared and distinct associations across SD-latent classes. *SPATA4* cg02833127 was consistently hypomethylated among cases in the AD+TD, CoD+TD, and PSU latent classes. This CpG site has been associated with aging trajectories from birth to late adolescence with methylation decreasing later in life ^55^. *SPATA4* locus has been also identified in a large-scale GWAS of educational attainment^40^. While the function of this locus is still unknown, a study observed aging-related changes in mice overexpressing this gene ^56^. In this context, *SPATA4* cg02833127 association across SD-latent classes points to the possible impact of SD comorbidities on aging-related regulatory mechanisms. We observed cross-ancestry heterogeneity in the association of this CpG site with AD+TD latent class with greater effect in EUR than AFR. This supports that SD comorbidities may affect biological pathways differently among population groups. Alternatively, cross-ancestry heterogeneity of the association between *SPATA4* cg02833127 and AD+TD latent class may reflect the contribution of environmental factors acting differently across population groups.

The AD+TD-latent class was uniquely associated with four additional CpG sites. Among them, *NOTCH1* cg13725899 was previously identified by multiple EWAS^57–60^. Two of which were conducted using brain tissues and linked this CpG site with fetal brain development ^58^and Alzheimer’s disease ^60^. Other *NOTCH1* CpG sites have been reported in EWAS of AD ^7^ and tobacco smoking ^61^. The *NOTCH1* locus was also identified in GWAS of brain-related outcomes, including cortical thickness and Alzheimer’s disease^62, 63^. The protein product of this gene plays an important role in a signaling pathway involved in neurodevelopmental processes^64^. *NOTCH1* may play a role in SD pathogenesis through its negative regulation of GABA receptors ^65^. Also, the other genes mapping to AD+TD-associated CpG sites (i.e., *ARID1B* and *SIN3B*) appear to play important regulatory roles in the central nervous system^66, 67^. Additionally, there were DNA methylation changes in *ARID1B* and *SIN3B* have been previously linked to cigarette smoking ^68–71^ and alcohol use ^72–74^.

The CoD+TD latent class showed specific epigenetic associations in *ANO6 and MOV10*. The latter locus has been linked to regulatory processes related to neuronal development and function^75^. Multiple GWAS identified *MOV10* in relation to cortical surface area, cortical thickness, and brain connectivity^76–78^. In contrast, *ANO6* gene does not appear to play a specific role in brain function, but is involved in phospholipid regulation occurring in various biological systems^79^. Previous EWAS identified other *ANO6* and *MOV10* CpG sites as associated with tobacco smoking ^80, 81^.

In addition to identifying individual CpG sites, we also observed a differentially methylated region mapping to *LDB1* that was associated with the PSU-latent class. This locus appears to be required for the early development of the dorsal telencephalon and the thalamus ^82^. Additionally, *LDB1* regulates gene expression in olfactory sensory neurons ^83^. The brain gene expression of this locus has been identified as negatively correlated with cocaine-seeking response in rats ^84^.

Overall, our EWAS findings support the association of SD-latent classes with genes involved in brain developmental processes. The applicability of our blood-based findings into brain mechanisms is supported by the high blood-brain correlation observed for several of the CpG sites identified (i.e., *MOV10* cg08355353, *SERTAD4* cg20270863, *SIN3B* cg06815056, *SPATA4* cg02833127, and *ARID1B* cg19436567). In some cases, genes we identified here (but with different CpG sites) were reported as associated by previous EWAS of tobacco smoking. Because we controlled for the effect of tobacco smoking on DNA methylation, the epigenetic associations identified in our study likely reflect SD comorbidities rather than the co-occurrence of tobacco smoking. Indeed, we did not identify any association between SD-latent classes and *AHRR* locus, which is the most established epigenetic biomarker for tobacco smoking ^85^.

We also identified genetic associations linking SD comorbidity patterns to other psychiatric disorders. PTSD was the only PGS showing Bonferroni significant association across the four SD-latent classes. This may support the role of mechanisms related to trauma response across SD comorbidity patterns. This is also in line with the well-known impact of stress response on biological mechanisms involved in SD pathogenesis^86^. TD-latent class was only associated with PTSD PGS. This may be due to the small sample size of the TD-latent class. Alternatively, it may also reflect the limited pleiotropy of this SD pattern with other mental illnesses. With respect to the other PGS associations shared among the remaining SD-latent classes, the relationships observed with respect to neurodevelopmental disorders (i.e., ADHD and schizophrenia) appear to converge with the EWAS findings, highlighting the potential role of altered neurodevelopmental processes in the shared pathogenesis among SD comorbidity patterns. Consistent with this hypothesis, previous studies highlighted how early-life adversity affects the development of brain reward and stress circuits increasing addiction vulnerability ^87^.

In addition to shared pleiotropic mechanisms, we also observed PGS associations specific to certain SD-latent classes. Bipolar-disorder PGS was uniquely associated with the AD+TD latent class. Multiple studies pointed to neural reward circuit dysfunction as the risk factor shared between bipolar disorder and SDs ^88^. The PSU-latent class showed two specific PGS associations: negative association with subjective well-being, and positive association with neuroticism. These may be related to poorer outcomes that are seen in individuals affected by the comorbidities of four SDs than those observed in subjects with only two comorbid SDs^89, 90^. In a previous study, a genetically inferred addiction factor shared across AD, TD, CaD, and TD was highly genetically correlated with neuroticism ^90^.

Our study has multiple limitations. The cohorts investigated were specifically recruited to study SD genetics, overrepresenting the samples for cases. This may reduce the generalizability of the SD-latent classes identified. The PGS analysis was limited to individuals of European descent, because of the lack of large-scale GWAS to derive statistically powerful PGS for other population groups. In the epigenome-wide analysis, we conducted a cross-ancestry meta-analysis assessing heterogeneity among the population groups. However, we had limited power to investigate ancestry-specific epigenetic effects. Similarly, we focused only on five main SDs because of the limited information on other substances in the cohorts investigated (e.g., hallucinogens, inhalants, sedatives, and amphetamines).

Notwithstanding these limitations, the present study provides new insights into genetic and epigenetic mechanisms contributing to in SD comorbidity patterns. Our findings support the potential role of brain developmental processes on SD pathogenesis and suggest possible mechanisms that differentiate the co-occurrence of different SD combinations. Building on this evidence, further studies are needed to extend these findings elucidating the dynamics underlying polysubstance use disorders.

## Data Availability

The data presented are included in the article and its supplemental material.

## ACKNOWLEDGEMENTS

This study is supported by the National Institute on Drug Abuse, R33 DA047527. GAP acknowledges support from the Yale Biological Sciences Training Program (T32 MH014276), Alzheimer’s Association (AARF-22-967171), NIH National Institute of Aging (K99AG078503), Yale Franke Fellowship in Science & Humanities, and Yale Women’s Faculty Forum Award. RP acknowledges grants from the National Institute of Mental Health (RF1 MH132337) and One Mind Rising Star Award. JDD acknowledges support by the National Institute on Drug Abuse K01 DA058807. DFL is funded by a Career Development Award from the US Department of Veterans Affairs Office of Research and Development (1IK2BX005058). HK acknowledges support from the Department of Veterans Affairs (VISN 4 MIRECC and I01 BX004820). JLMO acknowledges support from U.S. Department of Veterans Affairs via 1IK2CX002095 and NIDA R21DA050160. JG reports support from the Department of Veterans Affairs (5IO1CX001849-04 and the VISN 1 New England MIRECC) and NIH/NIDA (2R01DA037974, 1R01DA058862-01).

