## Supplemental Figures for "Epigenetic and Genetic Profiling of Comorbidity Patterns among Substance Dependence Diagnoses"

### **Supplementary Figures**

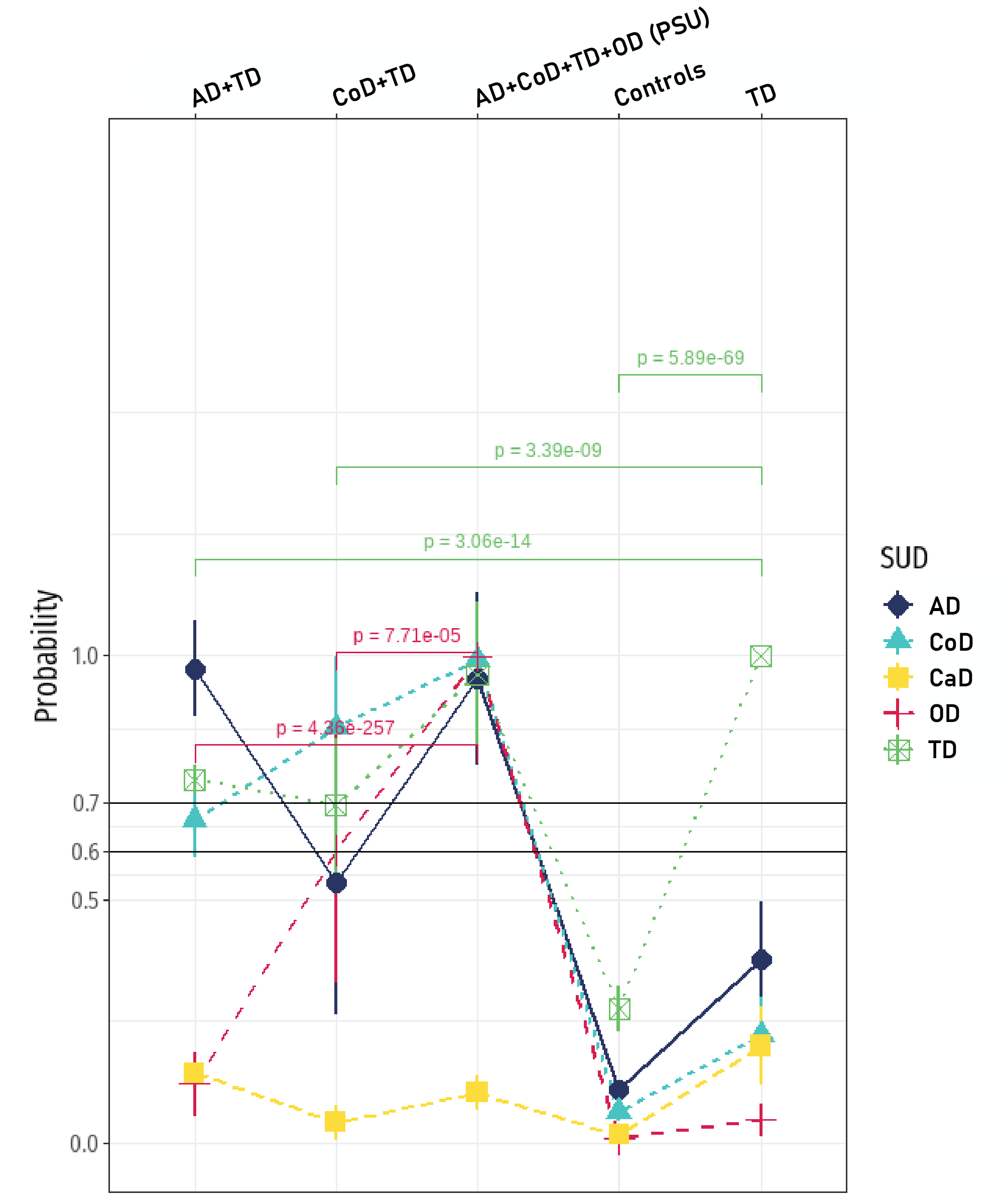

#### **Supplementary Figure 1.** The line graph shows statistical differences in probability value (y-axis) for SD cases across each latent class (x-axis: top). Each SD case group is shown in points (AD alcohol dependence – purple; CoD cocaine dependence – blue; CaD cannabis dependence – yellow; OD opioid dependence – pink; TD tobacco dependence – green).

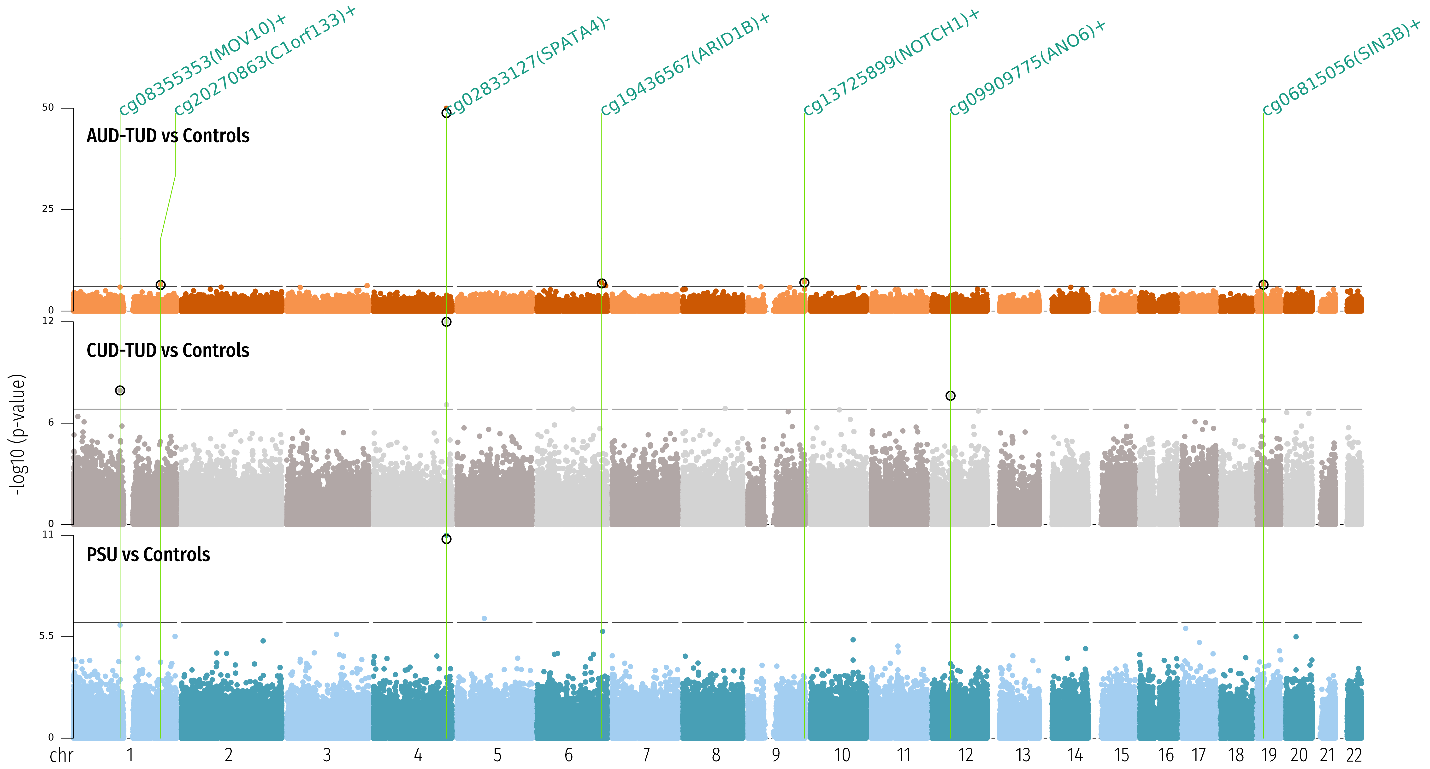

**Supplementary Figure 2. Manhattan Plot** showing differentially methylated probes in SUD latent classes versus Control class (class4) meta-analyzed from three ancestral cohorts – European (EUR), African (AFR), and Admixed/LatinX (AMR).

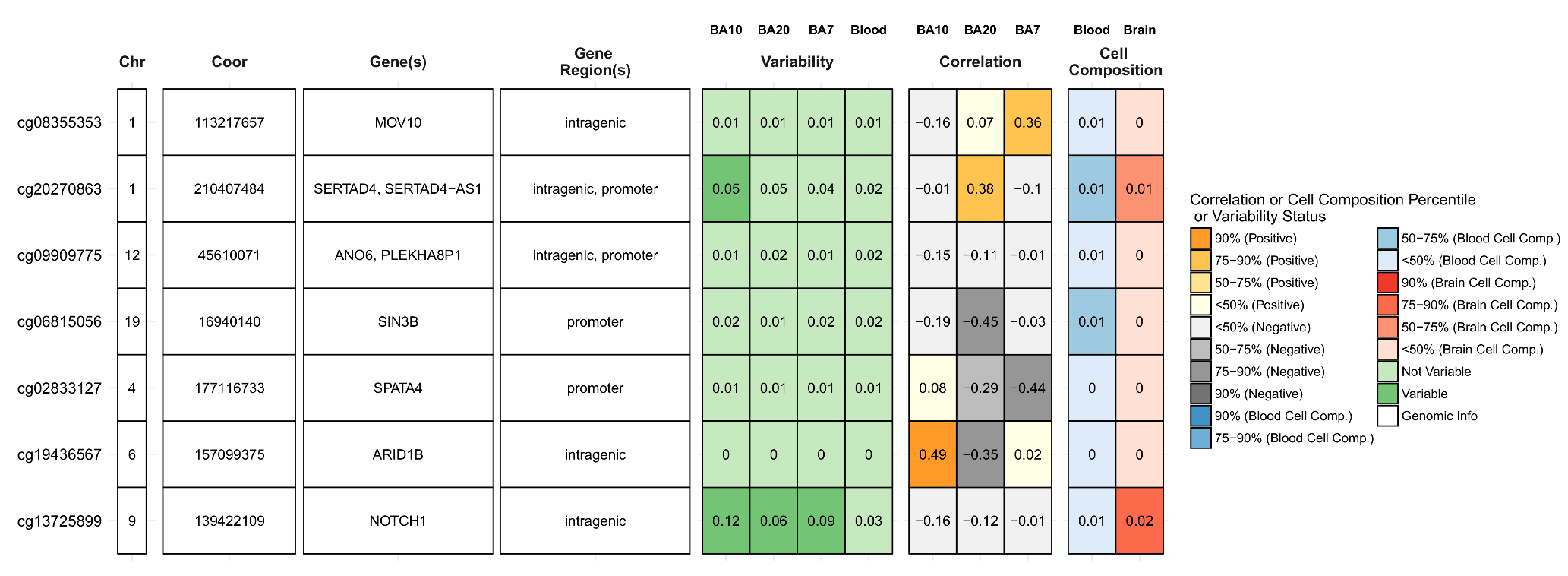

**Supplementary Figure 3.** Brain-blood methylation correlation of CpG sites identified in the epigenome-wide association study of SD-latent classes.
